# Association of Alcohol Intake with the Incidence of Atrial Fibrillation in Persons ≥65 Years Old in the Atherosclerosis Risk in Communities (ARIC) Cohort

**DOI:** 10.1101/2023.11.30.23299255

**Authors:** Louis Y Li, Linzi Li, Lin Yee Chen, Elsayed Z Soliman, Alvaro Alonso

## Abstract

**Background:** The association of alcohol intake with incident atrial fibrillation (AF) remains controversial, particularly among older individuals at higher risk of AF. This study explores the association of alcohol intake with incident AF in persons ≥65 years old in the Atherosclerosis Risk in Communities (ARIC) cohort.

**Methods:** Data were obtained from ARIC, a community-based cohort aiming to identify risk factors for cardiovascular disease. Alcohol intake was assessed through interviewer-administered questionnaires. Incident AF was ascertained between the 2011-2013 visit and 2019. Participants were classified as current, former, or never drinkers. Former drinkers were further categorized on weekly alcohol consumption: light, moderate, heavy. Covariates included demographic characteristics, prevalent cardiovascular disease, and other risk factors. The association between drinking characteristics and incident AF was analyzed using Cox proportional hazard models.

**Results:** There were 5,131 participants with mean (SD) age 75.2 (5.0) years, 41% male, 23% Black, and 739 (14%) cases of incident AF. Current (HR 1.04, 95% CI 0.83-1.29) and former (HR 1.16, 95% CI 0.93-1.45) drinkers had a similar risk of incident AF compared to never drinkers. Incident AF was similar across categories of alcohol consumption in former drinkers (heavy drinkers [HR 1.14, 95% CI 0.84-1.55], moderate drinkers [HR 1.15, 95% CI 0.75-1.78) compared to light drinkers). Risk of AF did not differ per 5-year increase in years of abstinence (HR 1.00, 95% CI 0.96-1.03). There was an increase in the risk per 5-year increase in years of drinking (HR 1.07, 95% CI 0.96-1.19).

**Conclusion:** We did not find consistent evidence supporting an increased risk of AF associated with alcohol intake in persons aged 65 and older, highlighting the need to further explore this relationship in older populations.

## Introduction

Atrial fibrillation (AF) is the most common cardiac arrhythmia, and its incidence and prevalence continue to increase worldwide.^1^ Alcohol intake has been proposed as a potential risk factor in AF incidence in both acute drinking settings and in long-term consumption.^2-4^ From a pathophysiological perspective, acute alcohol consumption has direct effects on cellular, autonomic, and electrophysiological functions involved in arrhythmogenicity.^5^ Long-term alcohol consumption has direct effects on left atrial substrate including left atrial remodeling, dilation, and fibrosis as well as indirect effects via interactions with other AF risk factors such as hypertension and cardiomyopathy.^6^ Additionally, alcohol intake may affect the autonomic nervous system, and it has been proposed that modulation of vagal and parasympathetic mechanisms may be involved in the pathogenesis of AF.^7^

Previous literature on the association between alcohol intake and incident AF has been less definitive, especially in considering levels of alcohol consumption.^8-10^ Furthermore, the association between previous alcohol consumption and incident AF such as in former drinkers has not been well-explored. In a previous analysis using data from the Atherosclerosis Risk in Communities (ARIC) study, Dixit et al.^11^ addressed the association of alcohol intake and incident AF in persons aged 45-64 years old and concluded that, among former drinkers, the number of years of drinking and the amount of alcohol consumed were associated with increased risk of AF. Among former drinkers, they found that a longer duration of abstinence was associated with a decreased risk of AF. Furthermore, previous literature addressing alcohol intake and AF in elderly populations is limited. Mukamal et al.^12^ studied older adults ≥65 years in the Cardiovascular Health Study (CHS) and concluded that current moderate alcohol consumption was not associated with risk of incident AF and that former drinking was associated with higher risk of incident AF compared to never drinkers.

This study aims to further investigate the understudied population of patients ≥65 years, who are at increased risk of incident AF. Additionally, we hope to better characterize the relationship between more detailed characteristics of alcohol consumption among former drinkers including amount of alcohol consumed, number of years of abstinence, and number of years of drinking. We hypothesize that former drinkers and current drinkers will have a higher risk of incident AF than non-drinkers. We hypothesize that, within former drinkers, the number of years of abstinence will be associated with decreased risk of incident AF and that the number of years of drinking and amount of alcohol consumed with be associated with increased risk of incident AF.

## Methods

### Study population

The ARIC study is a community-based cohort study that has been ongoing since 1987 to identify risk factors for atherosclerosis and cardiovascular disease. The study recruited individuals aged 45-64 years from 4 field centers: Washington County, MD; Forsyth County, NC; Jackson, MS; and selected suburbs of Minneapolis, MN. In total, 15,792 participants were enrolled at Visit 1 between 1987-1989 which included examinations for medical, social, and demographic data. Participants underwent follow-up examination periodically at designated intervals and were surveilled for cardiovascular outcomes. Visit 5 examinations took place between 2011-2013 and involved 6,538 participants which served as the baseline for this study. Due to low sample sizes, two groups were excluded: (1) non-White and non-Black participants and (2) non-White participants from the Minneapolis and Washington County centers. Additional exclusion criteria included participants <65 years, missing information on alcohol consumption history at Visit 5, and prevalent AF. Prevalent AF was ascertained via ECGs performed at study visits and hospital discharge codes (ICD-9-CM: 427.3x; ICD-10-CM: I48.x) prior to Visit 5. Patients with missing covariates were also excluded, except for smoking status due to >10% of patients with missing smoking status. Statistical analysis was conducted for 5,131 participants that met study criteria.

### Assessment of incident atrial fibrillation

Incident AF was defined as any AF occurring after the participant’s Visit 5 examination (2011-2013) and before December 31, 2019 for 3 centers (Washington County, Forsyth County, Minneapolis suburbs) and before December 31, 2017 for Jackson, MS. Incident AF was ascertained via hospital discharge codes (ICD-9-CM: 427.3x; ICD-10-CM: I48.x) not occurring in the context of open cardiac surgery and from death certificates with AF as an underlying or contributing cause of death (ICD-10: I48).^13^

### Assessment of alcohol intake

Alcohol intake was assessed from the ARIC Smoking and Alcohol Use Form at Visit 5. Participants were asked the following: (1) if they had ever consumed alcoholic beverages, (2) if they presently drink alcoholic beverages, (3) approximately how many years ago they stopped drinking, and (4) how much and what type of alcohol they consumed daily and weekly. Participants were stratified according to their drinker status: current, former, and never drinker. For former drinkers, amount of alcohol consumed weekly, number of years of abstinence, and number of years of drinking were assessed at the participants’ most recent visit for which data was available. Within former drinkers, amount of alcohol consumed weekly was calculated assuming the following alcohol content: 4oz wine (10.8g), 12oz beer (13.2g), 1.5oz hard liquor (15.1g).^11,14^ Weekly alcohol consumption was stratified into 3 categories based on definitions provided by the CDC’s National Center for Health Statistics:^15^

- Light drinker: ≤3 drinks per week
- Moderate drinker: >3 drinks but ≤7 drinks per week for women; >3 drinks but ≤14 drinks per week for men
- Heavy drinker: >7 drinks per week for women; >14 drinks per week for men

### Assessment of additional covariates

Age, sex, race, education level, prevalent cardiovascular disease [coronary artery disease (CAD), heart failure (HF), and stroke], hypertension (HTN), HDL-C, LDL-C, use of antihypertensive medications, use of anticoagulants, diabetes, smoking status, and body mass index (BMI) were considered as potential confounders. Sex, race, and education level were obtained through self-report at Visit 1. Education level was stratified into 5 categories: high school or less, high school graduate, vocational school, college, and graduate/professional school.

Prevalent CAD, HF, stroke, hypertension, HDL-C, LDL-C, use of antihypertensive medications, use of anticoagulants, diabetes, smoking history, and BMI were assessed during Visit 5 examinations. Prevalent CAD was defined by myocardial infarction (MI) indicated on baseline ECG, self-reported MI, or cardiac procedure. Prevalent HF was determined by criteria from the ARIC heart failure research committee: any adjudicated HF event, any first position ICD-9 code of 428.x before 2005, any physician report of HF, two subsequent instances of self-reported HF or HF medication use, or an instance of self-reported HF or HF medication use with an elevated NT-proBNP value >125 pg/mL from Visit 4 or Visit 5. Prevalent stroke was defined by self-report of stroke. Hypertension was defined as systolic blood pressure ≥140 mmHg, diastolic blood pressure ≥90 mmHg, or use of antihypertensive medications. HDL-C and LDL-C (mg/dL) were obtained from laboratory analytes. Use of antihypertensive medications and anticoagulants were obtained through self-report of use in the past 4 weeks. Diabetes was defined as fasting glucose ≥126 mg/dL, non-fasting glucose ≥200 mg/dL, current use of diabetes medication, or self-reported physician diagnosis of diabetes. Smoking status was stratified into 3 categories: current, former, and never smoker. BMI (kg/m^2^) was calculated from height and weight measurements.

### Statistical analysis

The association between drinker status and incident AF was analyzed using Cox proportional hazard models with adjustments for potential confounders. The proportional hazards assumption was assessed using Kaplan-Meier curves and log-minus-log survival plots for each covariate. Within former drinkers, the associations between number of years of drinking, number of years of abstinence, and amount of alcohol consumed weekly and incident AF were analyzed using Cox proportional hazard models. The number of years of abstinence and years of drinking were analyzed as continuous variables. Additionally, for years of abstinence, participants were stratified into quartiles and 20-year intervals; for years of drinking, participants were stratified into quartiles and 10-year intervals. Statistical analysis was conducted in SAS software (Version 9.4; SAS Institute, Cary, NC, US).

## Results

### Demographic characteristics

There were 5,131 participants included in the analysis (Figure 1). The mean (SD) age of participants was 75.2 (5.0) years, 41% were male, and 23% were Black. Of the 5,131 participants, there were 2,544 (50%) current drinkers, 1,475 (29%) former drinkers, and 1,112 (22%) never drinkers. Proportion of males was higher in current and former drinkers compared to never drinkers (47%, 45%, 21%, respectively). Lastly, never drinkers were more frequently likely to be never smokers compared to current and former drinkers (67%, 32%, 32%, respectively).

**Figure 1.**
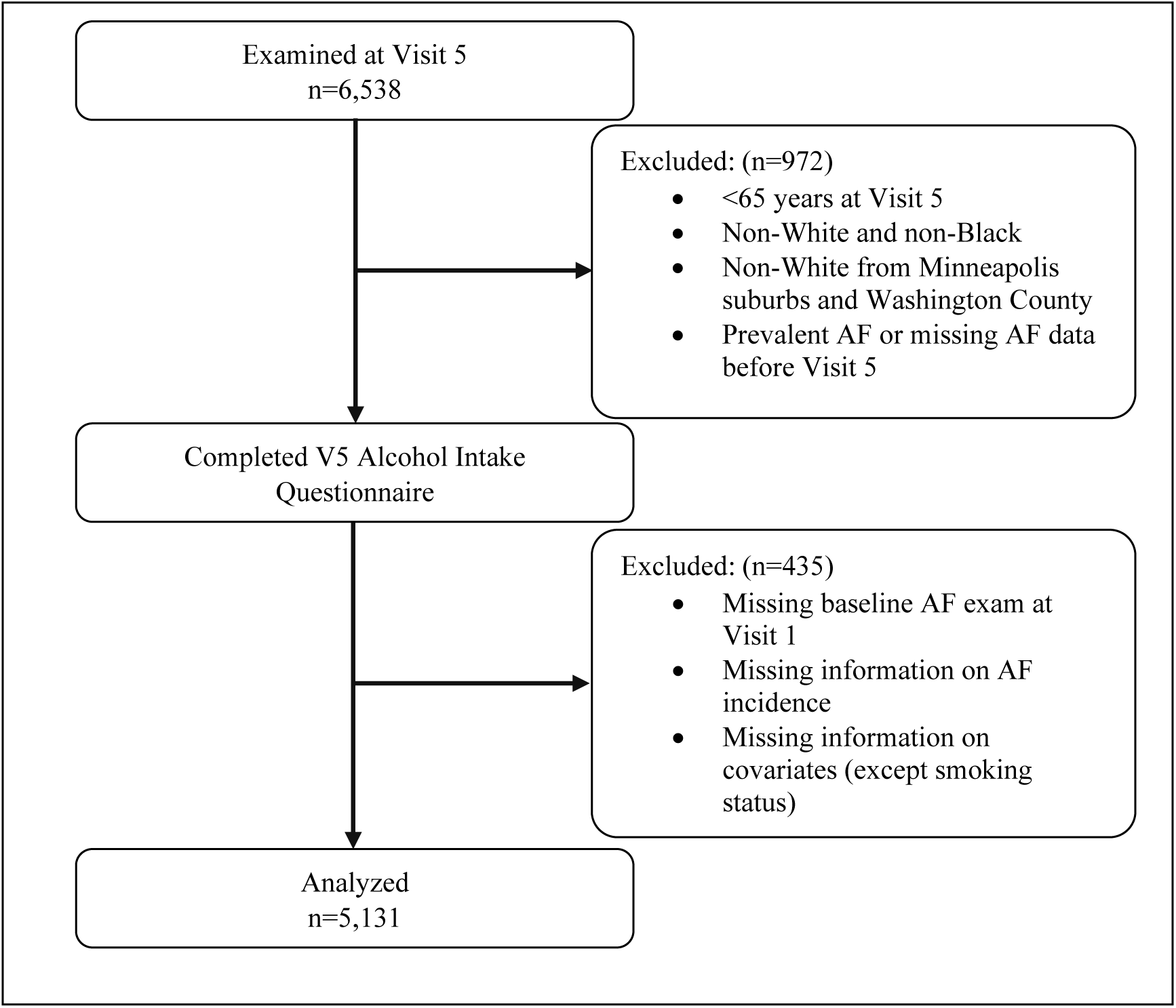
Study population flowchart.

### AF incidence

In total, 739 (14%) participants had incident AF: 377 (15%) current drinkers, 226 (15%) former drinkers, and 136 (12%) never drinkers. There was not a statistically significant difference in the risk of incident AF between categories (Figure 2). Participants were followed for a median time of 7.0 years. The overall incidence of AF was 23.2 cases per 1,000 person-years (PY): current drinkers (23.0 cases per 1,000 PYs), former drinkers (25.8 cases per 1,000 PYs), never drinkers (20.4 cases per 1,000 PYs).

**Figure 2.**
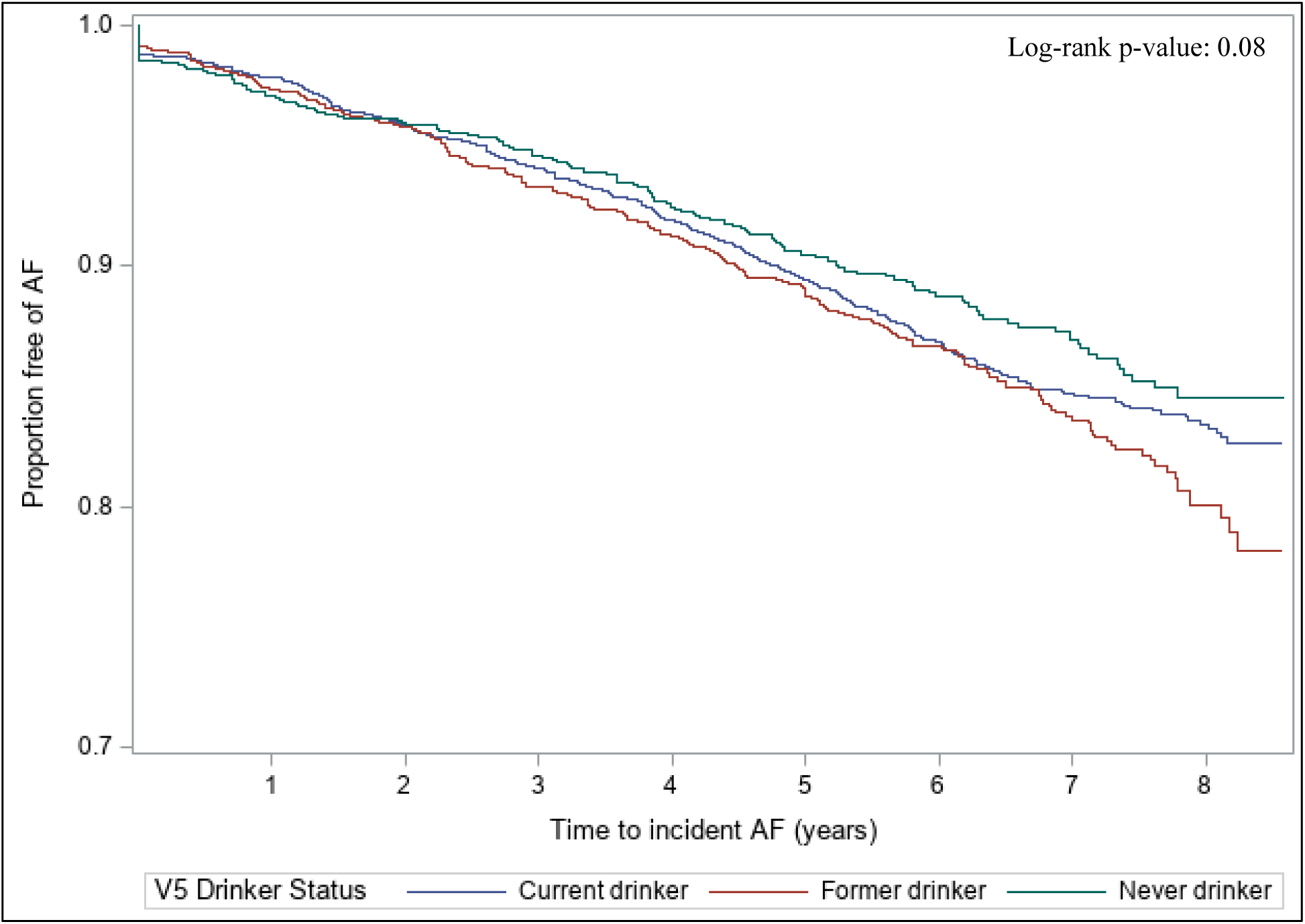
Kaplan-Meier curves for incident atrial fibrillation by drinker status.

### Drinker status

In unadjusted analyses, current drinkers had a higher risk of incident AF compared to never drinkers; however, the result was not statistically significant (HR 1.12, 95% CI 0.92-1.37). Former drinkers had a higher risk of incident AF compared to never drinkers (HR 1.27, 95% CI 1.02-1.57, p-value 0.03). After adjusting for covariates, the risk was attenuated in both current and former drinkers, and the hazard ratios were not statistically significant (HR 1.04, 95% CI 0.83-1.29; HR 1.16, 95% CI 0.93-1.45, respectively) (Table 2).

**Table 1.**
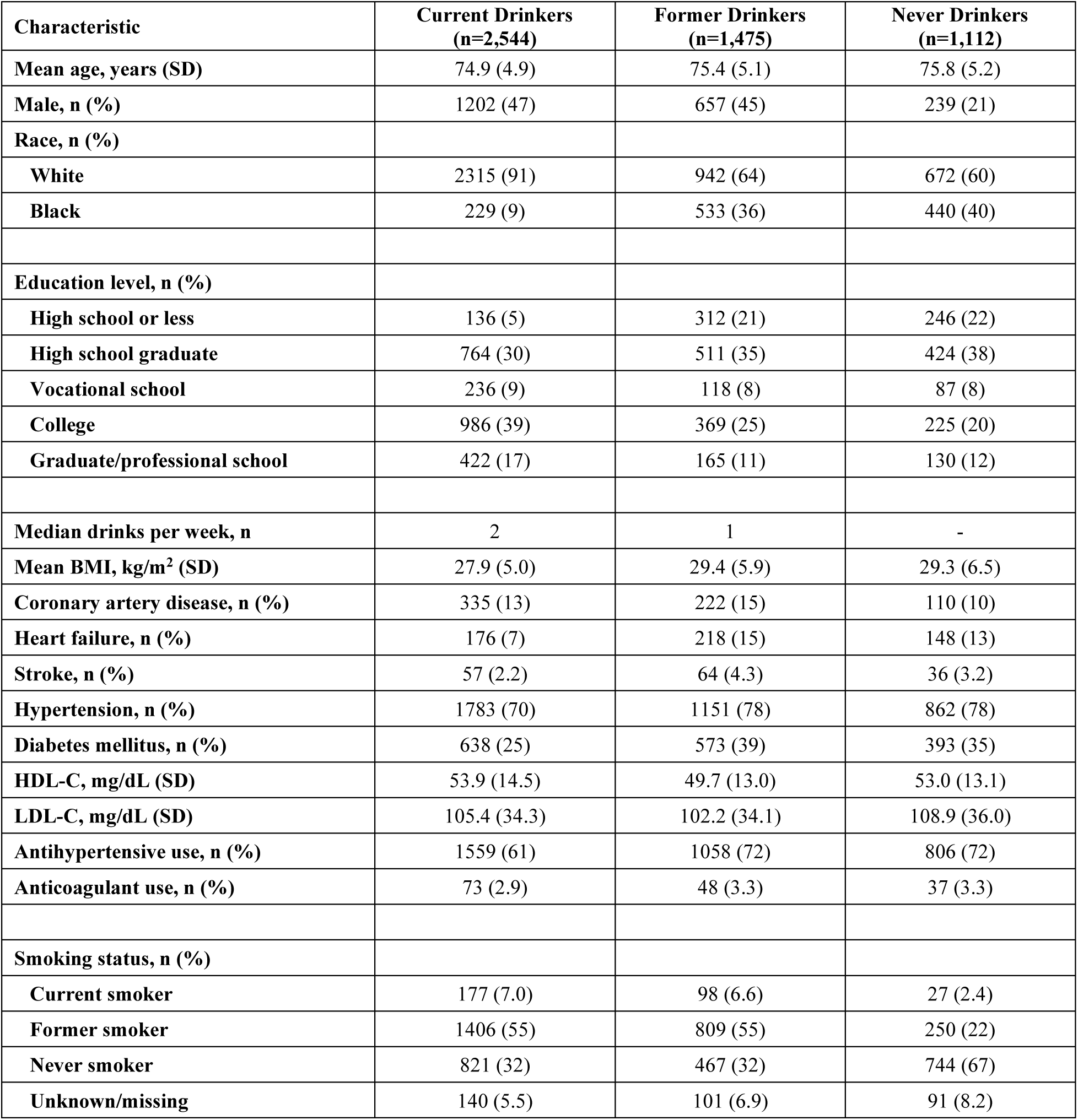
Demographic characteristics (n=5,131)

**Table 2.** Risk of incident atrial fibrillation by drinker status (n=5,131)

|  | AF Cases | Incidence Rate<br>(per 1,000 PYs) | Unadjusted<br>Hazard Ratio | 95% Confidence<br>Interval | p-value | Adjusted<br>Hazard Ratio* | 95% Confidence<br>Interval | p-value |
| --- | --- | --- | --- | --- | --- | --- | --- | --- |
| **Never** | 136 (12%) | 20.4 | 1 (Ref.) |  |  | 1 (Ref.) |  |  |
| **Current** | 377 (15%) | 23.0 | 1.12 | 0.92-1.37 | 0.25 | 1.04 | 0.83-1.29 | 0.75 |
| **Former** | 226 (15%) | 25.8 | 1.27 | 1.02-1.57 | 0.03 | 1.16 | 0.93-1.45 | 0.20 |

\*Adjusted for age, sex, race, education level, prevalent cardiovascular disease [coronary artery disease (CAD), heart failure (HF), and stroke], hypertension (HTN), HDL-C, LDL-C, use of antihypertensive medications, use of anticoagulants, diabetes, smoking status, and body mass index (BMI).

### Former drinkers

Within former drinkers, 1,431 out of 1,475 participants had data available on the number of drinks consumed weekly and were stratified into 3 categories: light, moderate, and heavy drinkers. There were 882 (62%) light drinkers, 163 (11%) moderate drinkers, and 386 (27%) heavy drinkers. There were 221 (15%) cases of incident AF: 128 (15%) occurring in light drinkers, 28 (17%) occurring in moderate drinkers, and 65 (17%) occurring in heavy drinkers. There was not a statistically significant difference in the risk of incident AF between categories (Figure 3). In unadjusted analyses, heavy and moderate drinkers had a higher risk of incident AF compared to light drinkers (HR 1.25, 95% CI 0.93-1.69; HR 1.22, 95% CI 0.81-1.83). In adjusted analyses, the comparisons were attenuated for both heavy (HR 1.14, 95% CI 0.84-1.55) and moderate (HR 1.15, 95% CI 0.75-1.78) drinkers (Table 3).

**Figure 3.**
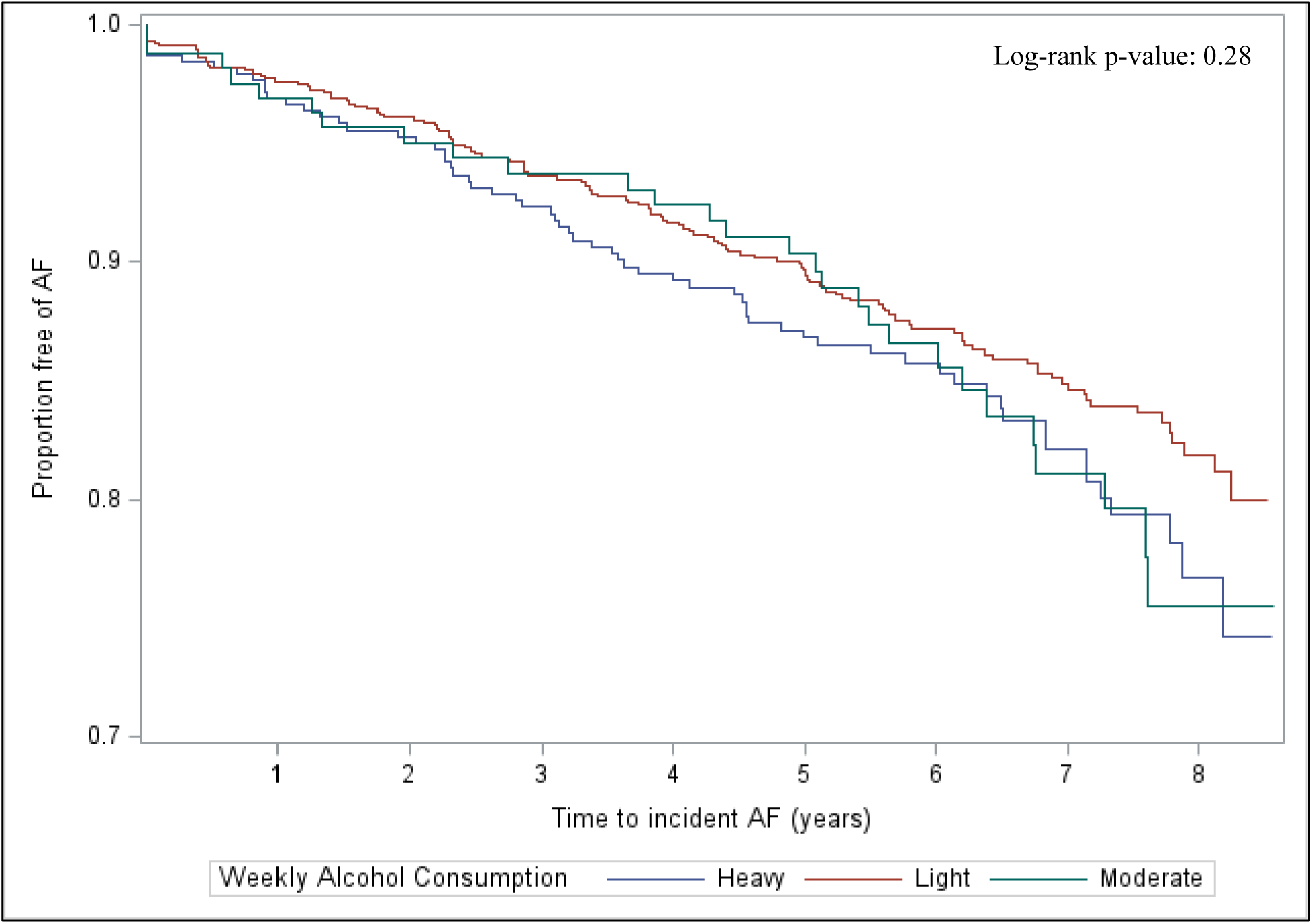
Kaplan-Meier curves for incident atrial fibrillation by weekly alcohol consumption in former drinkers.

**Table 3.** Risk of incident atrial fibrillation by weekly alcohol consumption in former drinkers (n=1,431)

|  | AF Cases | Incidence Rate<br>(per 1,000 PYs) | Unadjusted<br>Hazard Ratio | 95% Confidence<br>Interval | p-value | Adjusted<br>Hazard Ratio* | 95% Confidence<br>Interval | p-value |
| --- | --- | --- | --- | --- | --- | --- | --- | --- |
| **Light** | 128 (15%) | 23.9 | 1 (Ref.) |  |  | 1 (Ref.) |  |  |
| **Moderate** | 28 (17%) | 28.0 | 1.22 | 0.81-1.83 | 0.35 | 1.15 | 0.75-1.78 | 0.53 |
| **Heavy** | 65 (17%) | 29.4 | 1.25 | 0.93-1.69 | 0.14 | 1.14 | 0.84-1.55 | 0.41 |

\*Adjusted for age, sex, race, education level, prevalent cardiovascular disease [coronary artery disease (CAD), heart failure (HF), and stroke], hypertension (HTN), HDL-C, LDL-C, use of antihypertensive medications, use of anticoagulants, diabetes, smoking status, and body mass index (BMI).

Incident AF in former drinkers was also analyzed on the number of years of abstinence and number of years of drinking for 1,393 and 676 participants, respectively, who had available data. The median (IQR) years of abstinence was 20 (30) years, the range was 0-74 years, and there were 214 (15%) cases of incident AF. The median (IQR) years of drinking was 10 (15) years, the range was 0-43 years, and there were 102 (15%) cases of incident AF. There was no difference in the risk of incident AF per 5 years of abstinence in unadjusted (HR 1.00, 95% CI 0.96-1.03) or adjusted (HR 0.98, 95% CI 0.94-1.01) analyses. There was a small increase in the risk of incident AF per 5 years of drinking in unadjusted (HR 1.10, 95% CI 0.99-1.22) that was attenuated in adjusted analysis (HR 1.07, 95% CI 0.96-1.19) (Table 4). In unadjusted and adjusted analyses by quartile and by 10-year and 20-year intervals did not yield statistically significant results for years of abstinence or years of drinking (Supplemental Tables 1-4).

**Table 4.** Risk of incident atrial fibrillation by years of abstinence and years of drinking in former drinkers (n=1,393; n=676)

|  | Unadjusted<br>Hazard Ratio | 95% Confidence<br>Interval | p-value | Adjusted<br>Hazard Ratio* | 95% Confidence<br>Interval | p-value |
| --- | --- | --- | --- | --- | --- | --- |
| **Years of abstinence** | 1.00† | 0.96-1.03 | 0.84 | 0.98† | 0.94-1.01 | 0.22 |
| **Years of drinking** | 1.10† | 0.99-1.22 | 0.07 | 1.07† | 0.96-1.19 | 0.24 |

\*Adjusted for age, sex, race, education level, prevalent cardiovascular disease [coronary artery disease (CAD), heart failure (HF), and stroke], hypertension (HTN), HDL-C, LDL-C, use of antihypertensive medications, use of anticoagulants, diabetes, smoking status, and body mass index (BMI).
†Hazard ratio per 5 years.

## Discussion

In a community-based cohort study exploring the association between past alcohol intake and incident AF in elderly participants ≥65 years, the overall incidence of AF was 23.2 cases per 1,000 person-years, which is consistent with previous literature estimating the incidence in persons ≥65 years between 14.9-28.3 per 1,000 person-years.^16-18^ Current drinkers had a 4% higher risk of incident AF compared to never drinkers and former drinkers had a 12% higher risk of incident AF compared to never drinkers in adjusted analysis; however, these results were not statistically significant. While we hypothesized that current drinkers to have a higher risk than former drinkers, a possible explanation could involve participants’ reasons for quitting drinking such as underlying health conditions that may be associated with AF pathogenesis. Few studies have focused on this relationship in elderly populations, and the results have been inconclusive. In the Cardiovascular Health Study (CHS), Mukamal et al.^12^ demonstrated that current moderate alcohol consumption was not associated with the risk of incident AF, but former drinkers were at higher risk than never drinkers. In an elderly Chinese population, Ye et al.^19^ concluded that current drinkers were at higher risk than never drinkers in women only and not men. The results from this study show weak associations between drinker status and incident AF and do not support a strong effect of alcohol intake on AF risk in people aged 65 and older. The relationship likely remains multifaceted and complex, possibly depending on alcohol drinking patterns, prevalence of other risk factors, underlying risk of AF, and methodological aspects including methods to assess alcohol consumption and incident AF, thus requiring further investigation into the direct mechanisms of alcohol consumption on AF pathogenesis.

Within former drinkers, moderate drinkers had a higher risk of incident AF compared to light drinkers, heavy drinkers had a higher risk compared to light drinkers, and heavy drinkers had a similar risk to moderate drinkers; however, these results were not statistically significant. Studies addressing more detailed amount of alcohol consumed suggest an association between increasing alcohol consumption and incident AF. In the Framingham Heart Study (FHS) with a cohort aged 28-62 years, there was little association between long-term moderate alcohol consumption and the risk of incident AF, but there was a significant association between heavy consumption and incident AF.^3^ A separate study conducted in the FHS on a middle-aged population determined that that every 10 g per day of additional alcohol consumed was associated with a 5% higher risk of incident AF.^6^ A previous analysis on the ARIC cohort addressed baseline alcohol intake and incident AF in a younger population aged 45-64 years. The results demonstrated that greater alcohol consumption was associated with an increased risk of incident AF in persons aged 45-64 years.^11^ Previous literature suggests that heavy alcohol consumption may increase the risk of incident AF in younger populations; however, the findings from this study are not conclusive in supporting this hypothesis in elderly populations.

In this study focusing on persons ≥65 years, years of abstinence were not associated with an increased risk of incident AF. Each 5-year increase in years of drinking was associated with a 7% increased risk of incident AF; however, this association was not statistically significant. The previous ARIC analysis by Dixit et al.^11^ demonstrated that a longer duration of alcohol abstinence among former drinkers was associated with a lower risk of incident AF and greater alcohol consumption was associated with a higher risk of incident AF in persons aged 45-64 years. The variable conclusions between this study and Dixit et al.^11^ may suggest a fundamental difference between the study populations because of age. In the younger population, alcohol consumption may be a greater risk factor for incident AF; however, in elderly populations, alcohol consumption may not play as large of a role in the development of AF and may be masked by other, more significant risk factors associated with aging. To better characterize these differences, future investigations may compare younger and older populations as well as identifying the specific pathophysiologic changes that occur in the context of alcohol consumption.

Strengths of this study include extensive follow-up time and the quality of assessments of alcohol intake and additional covariates. Furthermore, to our knowledge, this study is only the second U.S.-based investigation in an elderly population on the association between alcohol consumption and incident AF. There were several limitations in this study. First, assessments of alcohol intake were reliant on self-report data through questionnaires. Self-report of alcohol intake may introduce recall bias but has been previously identified as a reliable and valid approach, and careful attention was given to accurately represent the data.^20,21^ Second, due to incident AF being ascertained from ICD codes, data on the severity and type of AF, such as paroxysmal vs. persistent vs. permanent AF was not captured, which may be an important distinction when considering the potential effects of alcohol consumption. Third, data was not collected on participants’ reasons for quitting drinking as health concerns may potentially explain the increased risk of incident AF among former drinkers. Last, while efforts were made to adjust for measured confounders, residual confounding may lead to bias in the reported results.

Our findings suggest that the association between alcohol intake and incident AF may be less conclusive and more multifaceted in elderly populations than in the general population. Alcohol intake may play a less important role in the pathogenesis of AF in elderly populations and is worth further exploration. Future investigations in elderly populations should compare them to general and younger populations, identify other potential risk factors that occur during the aging process that may be more involved in AF pathogenesis and may mask or supersede the effects of alcohol intake, and address concerns regarding the severity and type of AF patients may experience. As the global burden of AF continues to rise, better understandings of AF are a necessary and worthwhile endeavor, especially in elderly populations who are at greatest risk.

## Data Availability

All data produced in the present study are available upon reasonable request to the authors and ARIC Committee

https://aric.cscc.unc.edu/aric9/

## Non-standard Abbreviations and Acronyms

(in order of appearance)

AF: atrial fibrillation
ARIC: Atherosclerosis Risk in Communities
CHS: Cardiovascular Health Study
CAD: coronary artery disease
HF: heart failure
HTN: hypertension
BMI: body mass index
MI: myocardial infarction
PYs: person-years
FHS: Framingham Heart Study

## Acknowledgments

The authors thank the staff and participants of the ARIC study for their important contributions.

## Supplemental Material

Tables S1-S4

