## Supplemental Tables 1-4 for "Association of Alcohol Intake with the Incidence of Atrial Fibrillation in Persons ≥65 Years Old in the Atherosclerosis Risk in Communities (ARIC) Cohort"

**Supplemental Table 1.** Risk of incident atrial fibrillation by quartiles of years of abstinence in former drinkers (n=1,393)

| | Unadjusted Hazard Ratio | 95% Confidence Interval | p-value | Adjusted Hazard Ratio* | 95% Confidence Interval | p-value |
|---|---|---|---|---|---|---|
| **Quartile 1 (0-8 yrs)** | 1 (Ref.) | | | 1 (Ref.) | | |
| **Quartile 2 (9-25 yrs)** | 1.30 | 0.92-1.84 | 0.14 | 1.33 | 0.93-1.90 | 0.12 |
| **Quartile 3 (26-38 yrs)** | 1.07 | 0.68-1.67 | 0.78 | 0.77 | 0.50-1.17 | 0.93 |
| **Quartile 4 (39-75 yrs)** | 1.00 | 0.68-1.48 | 0.99 | 0.67 | 0.47-0.96 | 0.57 |

*Adjusted for age, sex, race, education level, prevalent cardiovascular disease [coronary artery disease (CAD), heart failure (HF), and stroke], hypertension (HTN), HDL-C, LDL-C, use of antihypertensive medications, use of anticoagulants, diabetes, smoking status, and body mass index (BMI).

**Supplemental Table 2.** Risk of incident atrial fibrillation by 20-year intervals of years in former drinkers (n=1,393)

| | Unadjusted Hazard Ratio | 95% Confidence Interval | p-value | Adjusted Hazard Ratio* | 95% Confidence Interval | p-value |
|---|---|---|---|---|---|---|
| **0-20 yrs** | 1 (Ref.) | | | 1 (Ref.) | | |
| **21-40 yrs** | 0.91 | 0.66-1.24 | 0.55 | 0.84 | 0.61-1.15 | 0.27 |
| **41-60 yrs** | 0.92 | 0.59-1.42 | 0.37 | 0.87 | 0.56-1.35 | 0.12 |
| **61-80 yrs** | 1.26 | 0.47-3.46 | 0.80 | 0.80 | 0.28-2.27 | 0.44 |

*Adjusted for age, sex, race, education level, prevalent cardiovascular disease [coronary artery disease (CAD), heart failure (HF), and stroke], hypertension (HTN), HDL-C, LDL-C, use of antihypertensive medications, use of anticoagulants, diabetes, smoking status, and body mass index (BMI).

**Supplemental Table 3.** Risk of incident atrial fibrillation by quartiles of years of drinking in former drinkers (n=676)

| | Unadjusted Hazard Ratio | 95% Confidence Interval | p-value | Adjusted Hazard Ratio* | 95% Confidence Interval | p-value |
|---|---|---|---|---|---|---|
| **Quartile 1 (0-5 yrs)** | 1 (Ref.) | | | 1 (Ref.) | | |
| **Quartile 2 (6-10 yrs)** | 1.59 | 0.78-3.27 | 0.20 | 1.67 | 0.80-3.48 | 0.17 |
| **Quartile 3 (11-20 yrs)** | 1.47 | 0.75-2.91 | 0.27 | 1.59 | 0.79-3.17 | 0.19 |
| **Quartile 4 (21-43 yrs)** | 1.21 | 0.6-2.36 | 0.58 | 1.19 | 0.60-2.35 | 0.62 |

*Adjusted for age, sex, race, education level, prevalent cardiovascular disease [coronary artery disease (CAD), heart failure (HF), and stroke], hypertension (HTN), HDL-C, LDL-C, use of antihypertensive medications, use of anticoagulants, diabetes, smoking status, and body mass index (BMI).

**Supplemental Table 4.** Risk of incident atrial fibrillation by 10-year intervals for years of drinking in former drinkers (n=676)

| | Unadjusted Hazard Ratio | 95% Confidence Interval | p-value | Adjusted Hazard Ratio* | 95% Confidence Interval | p-value |
|---|---|---|---|---|---|---|
| **0-10 yrs** | 1 (Ref.) | | | 1 (Ref.) | | |
| **11-20 yrs** | 0.94 | 0.56-1.58 | 0.81 | 0.88 | 0.52-1.51 | 0.65 |
| **21-30 yrs** | 1.49 | 0.85-2.60 | 0.16 | 1.19 | 0.65-2.16 | 0.57 |
| **31-43 yrs†** | 1.93 | 0.90-4.14 | 0.09 | 1.86 | 0.81-4.26 | 0.14 |

*Adjusted for age, sex, race, education level, prevalent cardiovascular disease [coronary artery disease (CAD), heart failure (HF), and stroke], hypertension (HTN), HDL-C, LDL-C, use of antihypertensive medications, use of anticoagulants, diabetes, smoking status, and body mass index (BMI).

†The category of 31-43 years includes 1 participant who had >40 years of drinking and was not separated into an additional category due to low sample size. All other participants had between 31-40 years of drinking.
